# Development and psychometric testing of EPAT-16: A short and valid measure for patient-centeredness from the patient’s perspective

**DOI:** 10.1101/2025.02.13.25322218

**Authors:** Eva Christalle, Levente Kriston, Stefan Zeh, Hannah Führes, Alica Schellhorn, Pola Hahlweg, Jördis Zill, Martin Härter, Isabelle Scholl

**Author notes:** Corresponding author information: Prof. Dr. Isabelle Scholl, Dipl.-Psych., Martinistr. 52, 20246 Hamburg, Germany.

## Abstract

**Background:** We previously developed the EPAT-64, a patient-reported experience measure designed to assess patient-centeredness (PC) across 16 dimensions of the validated integrative model of PC. While its modular structure makes it highly adaptable to examine certain dimensions of PC, generating an overall PC score requires assessing all 64 items. This is often challenging in routine settings with limited resources. Therefore, we developed and psychometrically tested a 16-item short form.

**Methods:** We conducted a cross-sectional study involving adult inpatients and outpatients receiving treatment for cardiovascular diseases, cancer, musculoskeletal disorders, and mental health conditions in Germany. To ensure comprehensive content coverage, we selected one item per dimension based on content relevance as well as item characteristics such as item difficulty and item-total correlation. We assessed the structural validity of a unidimensional model using confirmatory factor analysis (CFA), measured reliability with McDonald’s Omega, and evaluated construct validity by investigating the intercorrelation of the EPAT-16 sum score with measures of general health status and satisfaction with care.

**Results:** All items of the final EPAT-16 showed high acceptability and good item-total correlations, with approximately two-thirds demonstrating appropriate item difficulty. CFA showed a slight misfit of the unidimensional model, but high average variance explained. McDonald’s Omega showed high reliability. All hypotheses about construct validity were confirmed.

**Conclusions:** The EPAT-16 demonstrated good psychometric properties, making it a feasible tool for assessing overall PC when resource constraints preclude using the full EPAT-64. In particular, it can be used in routine care for feedback and quality improvement, as well as in research to assess relationships with other relevant variables. Since its items were designed generically, it can be used for different medical conditions and settings, for example, for public reporting. Future research should evaluate the EPAT-16 in diverse, independent patient samples to confirm that its positive characteristics are consistent across different populations and cultures.

**Patient or Public Contribution:** Patients did not participate as active members of the research team. However, for the research proposal, several patient organizations provided positive feedback on the study aims and their relevance, signed a collaboration agreement, and later supported recruitment by distributing study information.

## INTRODUCTION

Robust measures for assessing the degree of patient-centeredness (PC) are essential for promoting and improving PC in healthcare settings.^1 2^ A commonly used approach to evaluate PC from the patient’s perspective is through patient-reported experience measures (PREMs), which focus on what or how patients experienced specific processes or behaviours in their healthcare.^3 4^ We developed the EPAT-64 questionnaire,^5^ designed to assess 16 dimensions of PC, based on the integrative model of PC (see Figure 1).^6 7^

**Figure 1:**
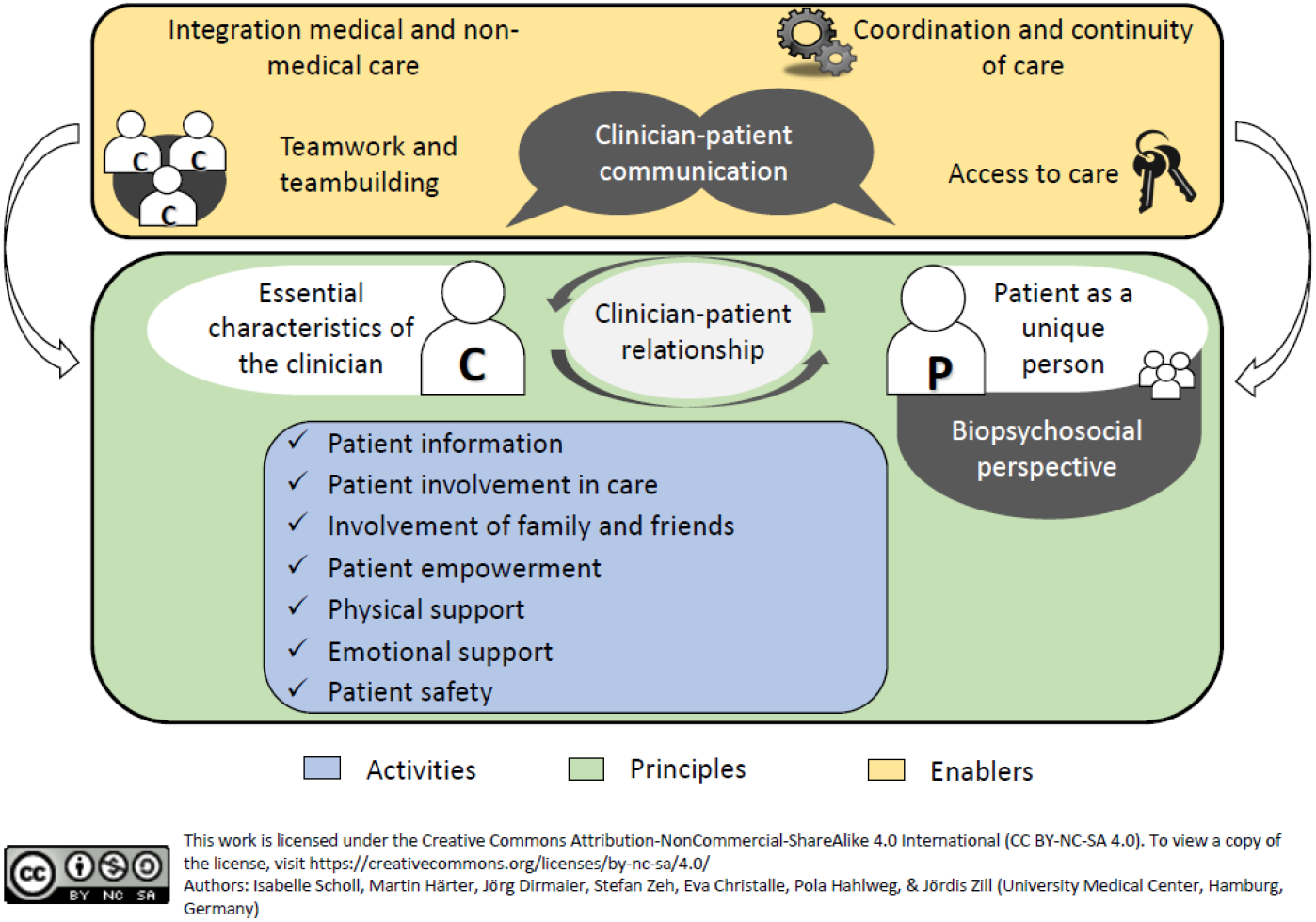
Integrative model of PC ^6 7^.

The development of the EPAT-64 followed a rigorous process, combining both theory- and data-driven approaches.^8^ It has demonstrated robust psychometric properties in a sample of 2,024 patients with chronic diseases.^5^ The EPAT-64 is available in both outpatient and inpatient versions, with each of the 16 dimensions represented by a module consisting of four items. Users can choose to use all 64 items or select specific modules that are most relevant to their objectives. While this modular approach ensures high adaptability, calculating an overall PC score requires administering all 64 items, which can be challenging. In particular for routine care — given constraints such as limited time, personnel, and financial resources — there is a need for a shorter version of the EPAT-64. Short scales can provide more efficient assessments in terms of time and cost.^9^ Additionally, shorter instruments reduce the burden on respondents,^10^ an important consideration for patients who may be cognitively impaired or fatigued due to treatments or their medical conditions. While longer measures offer greater content coverage, they can lead to higher dropout rates, thus compromising the representativeness of the sample.^11^ Reducing the number of items usually comes at the cost of reducing reliability, thus making short forms less applicable for individual level decision making. However, PC is usually assessed for a whole unit of care with feedback from several patients, which increases the reliability of the average score. Further, short forms are useful tools for multivariate studies, which examine a high number of variables and thus need to assess each variable with concise questionnaires to reduce burden on participants. Furthermore, short forms are in general useful tools for research questions focused on correlational relationships.^11^

The 16 dimensions of PC of the integrative model are considered to be interrelated.^6^ Psychometric analyses of the EPAT-64 revealed that the measure can be effectively represented by a bifactor model, which includes a general factor loading on all items as well as 16 specific factors representing each dimension.^5^ In this model, the general factor showed a strong correlation with treatment satisfaction, suggesting it may reflect the more subjective aspects of patient experiences. While the general factor was independent of the specific dimensions, the dimensions themselves were interrelated. Consequently, both theoretical and empirical evidence support the feasibility of deriving an overall PC score. However, it is important to note that such a score would encompass a combination of both the general factor and the specific factors. To preserve the breadth of the construct PC, each of the 16 dimensions should be represented in the short form.^12^

In this paper, we describe the development process of the EPAT-16, a shortened version of the EPAT-64, which evaluates each of the 16 dimensions using a single item. We present the psychometric properties of the EPAT-16 based on the sample used for item selection. Our goal was to design a tool to assess overall PC that can be used for all patients in routine care (e.g. as tool for continuous feedback or quality improvement), in research (e.g. to examine the relationship of a high number of constructs), and to compare different healthcare institutions regarding their overall level of PC.

## METHODS

### Study design

The EPAT-64 and its short form, the EPAT-16, were developed and tested as part of the ASPIRED study (Assessment of Patient-Centeredness Through Patient-Reported Experience Measures),^13^ a six-year mixed-methods research project on the measurement of patients’ perspectives on PC. Here, we report on the item selection process and psychometric evaluation of the EPAT-16, using the same dataset that was used for the development and evaluation of the EPAT-64.^5^ Since no specific guidelines exist for reporting psychometric evaluations of PREMs, we followed the COSMIN guidelines for reporting patient-reported outcome measures (PROMs) where applicable (see Appendix 1).

### The EPAT questionnaire

For psychometric testing, we conducted a cross-sectional survey that included all preliminary EPAT items. We developed these items based on the integrative model of PC^6 7^ shown in Figure 1, informed by a literature review, key informant interviews with experts, and focus groups with patients with chronic diseases. The preliminary questionnaire assessed 16 dimensions of PC, comprising 121 items for inpatients and 120 items for outpatients. Our goal was to create a generic instrument that would be applicable across different diseases. Participants rated the EPAT items on a 6-point Likert scale of agreement (1 = completely disagree, 2 = strongly disagree, 3 = somewhat disagree, 4 = somewhat agree, 5 = strongly agree, 6 = completely agree), with an additional “does not concern me” option for each item. Higher scores indicated a greater degree of PC, except for nine items, which we reverse-coded before analysis. We originally developed and tested the EPAT in German. For this paper, we translated the EPAT-16 into English using the TRAPD process, which involves a team-based approach consisting of translation, review, adjudication, pretesting, and documentation.^14^

### Data collection and participants

We used the same sample for the EPAT-16 item selection that we had used for the EPAT-64. We collected data through both paper-pencil questionnaires and an online survey conducted via LimeSurvey (LimeSurvey GmbH, Hamburg, Germany). For the paper-pencil data collection, we used consecutive sampling at 10 inpatient and 14 outpatient healthcare institutions (e.g., hospitals, primary care centers) in the metropolitan area of Hamburg, Germany, from June 2020 to February 2022. Outpatients received the questionnaire before their appointment and were asked to report on their experiences in the past four weeks. Inpatients received the questionnaire shortly before discharge and were asked to report on their entire inpatient stay. For the online survey, we recruited participants using community-based strategies, including social media and patient organizations from across Germany. Online participants were asked to refer to their last outpatient visit or inpatient stay within the past month. We included adults (18 years or older) with at least one of the following four medical conditions: cardiovascular diseases, cancer, musculoskeletal diseases, and mental disorders. Inclusion was based on self-report. As outlined in our study protocol, we aimed for a sample size of 250 participants in each of the two settings (outpatient and inpatient) for each of the four medical conditions, resulting in a target sample size of 2,000 participants.^13^

The full questionnaire for data collection included the preliminary version of the EPAT, the first item of the German version of the 12-item Short Form Survey,^15^ the German version of the 8-item Client Satisfaction Questionnaire (ZUF-8),^16^ the European Health Literacy Questionnaire (HLS-EU-Q16)^17^ in German as well as questions on demographic characteristics and health status. We also added two control items (e.g., “To show that you are reading attentively, please check ‘somewhat disagree’/’somewhat agree’”). Data collection was anonymous. Participants gave explicit informed consent (either written before receiving the paper-pencil questionnaire or per active click at the start of the online-survey) and could opt to receive a €10 incentive for completing the questionnaire.

### Data analyses

### Item characteristics and item selection strategy

We selected the items for the EPAT-16 during the same team discussion session in which we chose the items for the EPAT-64.^5^ Given that the original instrument assesses 16 dimensions, it has a rather broad scope. Even though the EPAT-16 omits those dimensions and instead aims to measure an overall degree of PC, it was important, that all dimensions are represented in the final short form to ensure content coverage.^12^ Further, it is recommended that item selection for short forms should be guided by both content of the items and item characteristics.^9^ Therefore, our selection strategy was to choose one item per dimension, focusing on how well the content represented the dimension’s core definition, but also considering the following item characteristics: (1) acceptance indicated by the percentage of missing responses,^18^ (2) relevance indicated by the percentage of ‘does not concern me’ replies, (3) item-difficulty indicated by the mean standardised to range from 0 to 1 (recommended range 0.2 to 0.8),^19^ (4) corrected item-total correlation within each dimension (recommended range >0.3),^20^. Additionally, in contrast to EPAT-64, we decided to create one version of EPAT-16 for both inpatient and outpatient settings. Hence, we excluded any items that applied to only one setting. This allowed us to create a short form that could be used across different healthcare contexts, providing comparability.

### Psychometric analyses of final EPAT-16

After item selection, we examined further psychometric properties of the EPAT-16 within the same data set. First, we examined structural validity using confirmatory factor analysis (CFA)^21-23^ testing a unidimensional model. We used a robust maximum likelihood estimator (MLR). As recommended,^25^ we used several indices to assess model fit (see Table 2). We report standardised factor loadings and average variance explained by calculating the average percentage of the variance in the items which is explained by the common factor. Further, based on the factor analysis, we calculated McDonald’s Omega to test reliability.^27^ We examined measure invariance between medical conditions by imposing the same factor structure (configural invariance), same factor loadings (weak invariance) and same factor loadings and intercepts (strong invariance) for each of the four groups.^28^ We compared fit of these models using likelihood ratio tests.

Next, we examined construct validity. To assess convergent validity, we hypothesized that the EPAT-16 would correlate with the German ZUF-8, a unidimensional measure of patient satisfaction with high internal consistency (Cronbach’s alpha = 0.9).^16^ We expected the correlation to exceed 0.5.^29^ For discriminant validity, we used the first item of the German SF-12, which assesses general health status, has an item difficulty of 0.4 and high acceptability.^15^ Given that health status is theoretically unrelated to PC, we hypothesized a correlation magnitude below 0.3.^29^ Finally, we examined the distribution of the EPAT-16 sum score using its mean, standard deviation, and percentage frequencies.

We used SPSS Version 27.0 (IBM Corp., Armonk, NY, USA) to enter data, calculate sample characteristics as well as sum scores and item total correlations of EPAT-16 and to test construct validity. All other data cleaning and analyses were performed using R Version 4.3.2 (R Core Team, Vienna, Austria). During data cleaning, we excluded patients who responded to fewer than 70% of the EPAT items (where “does not concern me” was counted as a response) or who failed both control items. We treated “does not concern me” responses as missing values in all further analyses. For factor analyses, we used full information maximum likelihood (FIML)^24^ to deal with missing values. For analysing construct validity, item-total-correlations and sum scores of EPAT-16, we used multiple imputation^33^ with fully conditional specification^34^ and predictive mean matching (40 imputations and maximum 100 iterations) to handle missing data.^33^ We analysed data of inpatient and outpatient settings separately, with results reported for all medical conditions combined.

## RESULTS

### Sample

Details of the sample were already reported in the primary publication on EPAT-64.^35^ The participant flow chart and the table with all sample characteristics are replicated in Appendix 2. The final analyses included n = 1,092 outpatient and n = 931 inpatient questionnaires. Of the outpatients, 59.1 % were female, 38.5 % male, and 0.5 % identified as diverse. Their mean age was 52.8 years (standard deviation (SD) = 17.4, range 18-95). Regarding medical conditions, 25.4 % were treated for cardiovascular diseases, 25.0 % for cancer, 19.9 % for musculoskeletal diseases, and 25.0% for mental disorders. In the inpatient sample, 41.3 % were female, 55.5 % male, and 0.6 % identified as diverse. The mean age was 55.5 years (SD = 17.4, range 18-92). Of the inpatients, 30.7 % were treated for cardiovascular diseases, 37.7 % for cancer, 9.9 % for musculoskeletal diseases, and 21.7 % for mental disorders.

### Item characteristics

The items selected for EPAT-16 are shown in Table 1 with their corresponding dimension of the EPAT-64, their English translation, and their item characteristics. None of the final EPAT-16 items had to be reversed.

Regarding relevance, in the outpatient sample three items had “does not concern me” response rates above 30% (highest rate was for the “physical support”-item with 54.7%). In the inpatient sample all items had “does not concern me” response rates below 30% (highest rate was for the item “involvement of family and friends” with 29.6%). Acceptance was good for all items (highest rate of missing responses was 1.4% for outpatients and 1.5% for inpatients). Regarding item difficulty, 12 and 11 items in the outpatient and inpatient sample, respectively, were within the recommended range. The remaining items showed ceiling effects. Item total correlations ranged from 0.403 to 0.748 in the outpatient sample and from 0.489 to 0.742 in the inpatient sample. Item characteristics per medical condition are shown in Appendix 3.

**Table 1:** Item characteristics of the EPAT-16.

| Dimension of EPAT-64 | Item | Outpatient sample |  |  |  |  |  |  | Inpatient sample |  |  |  |  |  |  |
| --- | --- | --- | --- | --- | --- | --- | --- | --- | --- | --- | --- | --- | --- | --- | --- |
|  |  | M | SD | does not concern me | No reply | Item difficulty | Item Total Corr | Std. factor loadings | M | SD | does not concern me | No reply | Item difficulty | Item Total Corr | Std. factor loadings |
| Essential characteristics of the clinician | The healthcare professionals were sensitive (for example they addressed my feelings, showed understanding, or empathized with my situation). | 5.1 | 1.2 | 4.2% | 1.2% | 0.82 | .714 | .816 | 5.1 | 1.2 | 3.9% | 0.8% | 0.82 | .701 | .798 |
| Clinician-patient relationship | I trusted my healthcare professionals. | 5.2 | 1.1 | 1.8% | 1.2% | 0.85 | .694 | .792 | 5.3 | 1.1 | 1.2% | 0.4% | 0.86 | .676 | .778 |
| Patient as a unique person | My wishes, needs and expectations were asked and taken into account in the treatment. | 4.8 | 1.4 | 8.3% | 0.8% | 0.75 | .748 | .829 | 4.8 | 1.3 | 5.5% | 0.9% | 0.76 | .742 | .803 |
| Biopsychosocial perspective | My entire personal life was taken into account during the treatment (for example, job, family and friends, partnership and sexuality, culture and religion, age, or financial circumstances). | 3.9 | 1.8 | 16.1% | 1.2% | 0.57 | .646 | .617 | 3.8 | 1.8 | 14.2% | 0.8% | 0.56 | .593 | .538 |
| Clinician-patient communication | I was given enough time to describe my concerns and my situation (for example, medical history or current symptoms) | 5.3 | 1.1 | 2.6% | 1.0% | 0.86 | .673 | .760 | 5.2 | 1.1 | 2.8% | 0.5% | 0.85 | .690 | .775 |
| Integration of medical and non-medical care | I was asked if I use or would like to use additional services (for example, support groups, counseling, health courses, complementary and alternative medicine, or spiritual support/pastoral care). | 3.1 | 2.0 | 30.4% | 1.1% | 0.42 | .493 | .439 | 3.4 | 2.0 | 25.8% | 1.5% | 0.49 | .531 | .464 |
| Teamwork and teambuilding | The processes within the team were well organized. | 5.1 | 1.0 | 7.1% | 0.8% | 0.83 | .403 | .407 | 5.1 | 1.1 | 1.2% | 1.0% | 0.82 | .605 | .695 |
| Access to care | If I wanted to speak to a physician, they were easily accessible. | 4.7 | 1.4 | 23.4% | 1.3% | 0.73 | .496 | .533 | 4.8 | 1.2 | 6.6% | 1.0% | 0.76 | .611 | .691 |
| Coordination and continuity of care | It was discussed with me whether follow-up appointments would be useful (for example, for aftercare or further treatment). | 5.1 | 1.4 | 13.1% | 1.4% | 0.82 | .479 | .498 | 5.0 | 1.4 | 7.2% | 1.0% | 0.80 | .547 | .582 |
| Patient safety | I was encouraged to speak up if I noticed inconsistencies in my treatment. | 3.9 | 1.8 | 23.4% | 0.8% | 0.58 | .614 | .601 | 4.1 | 1.7 | 14.2% | 1.2% | 0.62 | .610 | .589 |
| Patient information | I received information about my condition from my healthcare professionals (for example, causes, symptoms, effects or course). | 4.6 | 1.6 | 10.9% | 0.7% | 0.71 | .630 | .647 | 4.6 | 1.5 | 6.4% | 1.3% | 0.72 | .606 | .622 |
| Patient involvement in care | I was an equal partner with my healthcare professionals (for example, in making decisions or sharing information). | 5.0 | 1.2 | 7.6% | 1.3% | 0.79 | .665 | .760 | 4.8 | 1.3 | 4.4% | 1.8% | 0.76 | .643 | .728 |
| Involvement of family and friends | I was informed about the options for involving my family members in the treatment (for example, accompanying to appointments, participating in conversations, or assisting with medication intake). | 3.1 | 2.0 | 42.5% | 1.3% | 0.41 | .553 | .514 | 3.3 | 1.9 | 29.6% | 1.0% | 0.46 | .578 | .495 |
| Patient empowerment | I was encouraged to improve my health by changing my behavior (for example, through diet, exercise, reducing tobacco or alcohol). | 4.0 | 1.8 | 26.0% | 0.8% | 0.61 | .592 | .562 | 4.0 | 1.7 | 25.8% | 0.5% | 0.61 | .492 | .444 |
| Physical support | When I had pain, I was helped quickly. | 4.8 | 1.4 | 54.7% | 1.0% | 0.76 | .551 | .624 | 5.4 | 1.0 | 19.4% | 0.8% | 0.87 | .492 | .599 |
| Emotional support | The healthcare professionals addressed my fears and concerns (for example, by showing understanding and providing encouragement). | 4.4 | 1.7 | 19.2% | 0.8% | 0.68 | .731 | .771 | 4.4 | 1.6 | 15.9% | 1.5% | 0.69 | .688 | .688 |

Notes: M = mean, SD = standard deviation, Item Total Corr = Corrected item total correlation with the other EPAT-16 items, Std. factor loadings = Standardised factor loadings in the unidimensional model

**Table 2:** Model fit, average variance explained, reliability and measure invariance of the unidimensional model.

|  | Chi square |  |  | RMSEA | SRMR | TLI | CFI | AIC | BIC |
| --- | --- | --- | --- | --- | --- | --- | --- | --- | --- |
| | $\chi^2$ | df | P value | | | | | | |
| **Recommendation** |  |  | >.05 | ≤ .06 | <.08 | ≥ .90 | ≥ .90 | Smallest value |  |
| **Outpatient sample** | 843 | 104 | <0.001 | .081 | .066 | .877 | .894 | 43910 | 44149 |
| Configural invariance | 1170 | 416 | <0.001 | .097 | .080 | .856 | .875 | 41279 | 42229 |
| Weak invariance | 1280 | 461 | <0.001 | .095 | .100 | .861 | .866 | 41299 | 42026 |
| Strong invariance | 1816 | 506 | <0.001 | .114 | .114 | .799 | .788 | 41744 | 42249 |
| **Inpatient sample** | 856 | 104 | <0.001 | .088 | .073 | .865 | .883 | 39789 | 40022 |
| Configural invariance | 1024 | 416 | <0.001 | .086 | .069 | .881 | .897 | 38345 | 39272 |
| Weak invariance | 1122 | 461 | <0.001 | .086 | .096 | .881 | .886 | 38353 | 39063 |
| Strong invariance | 1796 | 506 | <0.001 | .115 | .122 | .790 | .778 | 38937 | 39429 |

Notes: Configural invariance = same factor structure imposed on all four groups, Weak invariance = Same factor loadings imposed on all four groups, Strong invariance = same factor loadings and intercepts imposed on all four groups

### Psychometric analyses of final EPAT-16

Standardised factor loadings are shown in Table 1. The ranged from 0.407 to 0.829 in the outpatient sample and from 0.444 to .803 in the inpatient sample. Except for SRMR, all model fit indices slightly differed from the recommended range (see Table 2). We further explored this misfit by calculating residual correlations between items and modification indices (see Appendix 4). The highest residual correlation was found between the items for “Integration of medical and non-medical care” and “Involvement of family and friends” (0.249 for outpatients and 0.278 for inpatients). The average variance explained of the unidimensional model was 94.7% for the outpatient sample and 94.5% for the inpatient sample. McDonald’s Omega indicated a high reliability of the unidimensional model (0.908 for the outpatient version and 0.906 for the inpatient version). Regarding measurement invariance the model fit indices exhibited only slight changes for configural invariance and weak invariance when compared to the CFA of the full dataset. For strong invariance, all model fit indices worsened noticeably. Yet, likelihood ratio tests for model comparisons (configural invariance vs. weak invariance, and weak invariance vs. strong invariance) showed a significant decline of fit (p < 0.001) for all comparisons in both samples.

Regarding construct validity, all hypotheses were confirmed. In the outpatient sample, the EPAT-16 sum score had a pooled correlation of 0.757 with satisfaction with care and of −0.113 with general health status. In the inpatient sample those correlations were 0.794 and −0.050 respectively.

EPAT-16 sum scores had mean = 72.6 (SD = 15.7) in the outpatient sample and mean = 73.6 (SD = 15.2) in the inpatient sample (maximum possible range is 16 (lowest degree of PC) to 96 (highest degree of PC). Distribution of EPAT-16 sum scores are shown in Figure 2. Mean, SD and percentiles for the EPAT-16 sum scores for the overall samples and per medical condition can be found in Appendix 5.

**Figure 2:**
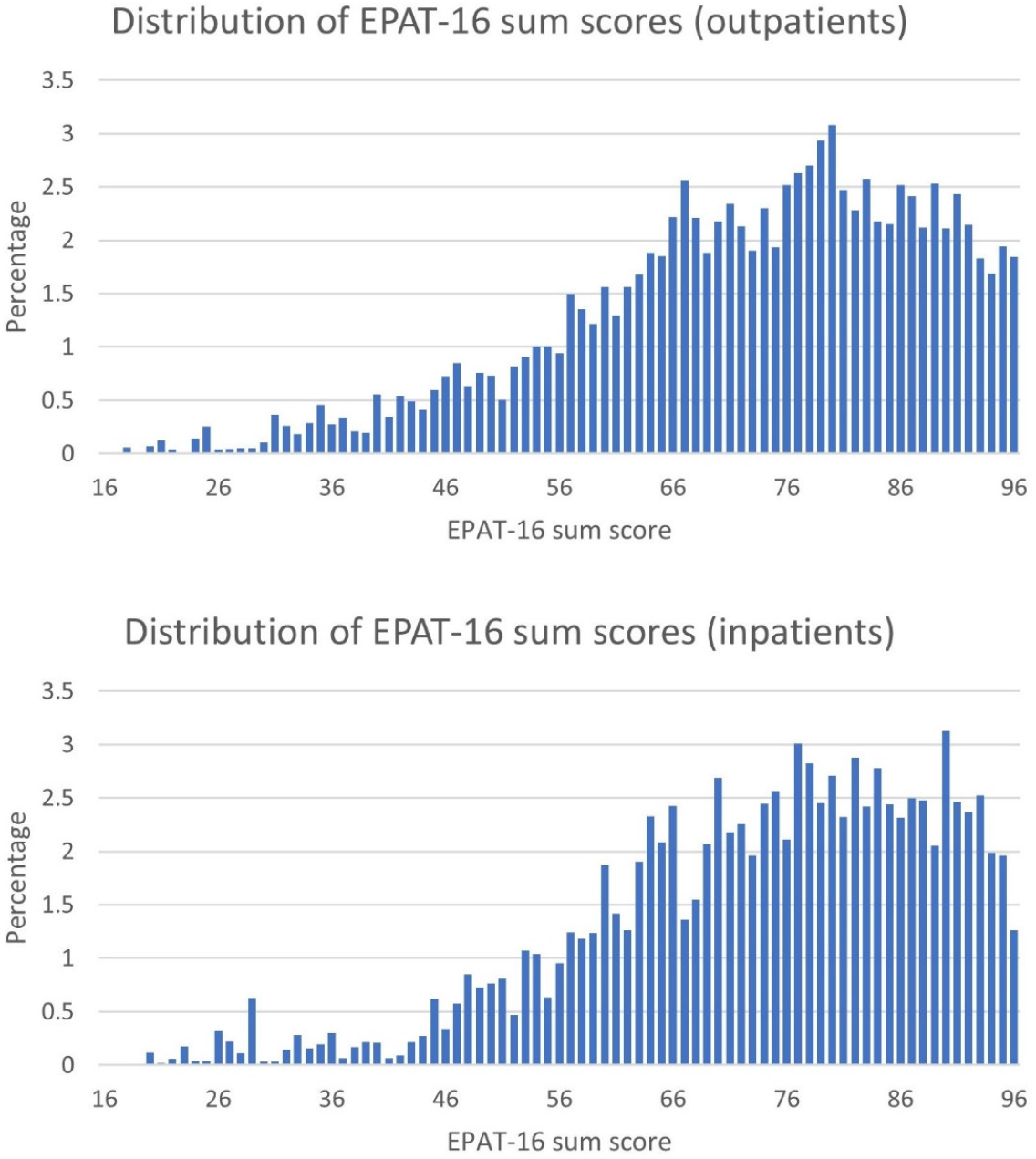
Percentage frequencies of EPAT-16 sum scores.

## DISCUSSION

### Summary

We reported on the development and item selection for the EPAT-16, a short form designed to assess the overall degree of PC. We also evaluated the psychometric properties of the final EPAT-16 within the item selection sample. All items of the EPAT-16 demonstrated high acceptability and good corrected item-total correlations. While all items showed high relevance in the inpatient sample, three items were marked as “does not concern me” by more than 30% of the outpatients. Approximately three-quarters of the items displayed appropriate difficulty levels, while the remaining items exhibited ceiling effects. The sum score showed a good degree of variance, although most scores were concentrated in the upper half of the possible range. Regarding structural validity, most model fit indices for the unidimensional model were slightly outside the recommended range. However, the model explained a substantial portion of the item variance, and McDonald’s Omega indicated high reliability. Even though the model fit indices changed only slightly when imposing same factor loadings, based on the likelihood ratio tests the EPAT-16 did not achieve weak measurement invariance. Finally, all hypotheses regarding construct validity were confirmed.

The EPAT-16 offers an efficient and concise method for assessing PC while ensuring representation of all 16 dimensions in the integrative model of PC.^6 7^ This was achieved by an item selection strategy that, next to item characteristics, took into account to which extent an item reflected the dimension definition. While the EPAT-64 allows to explore detailed questions about specific dimensions of PC, the EPAT-16 serves as a potentially valuable tool when the focus is on an overall PC score and resources are constrained. Reducing the number of items to a quarter makes the EPAT-16 more feasible in routine care, where time and personnel are limited, and patients might have low cognitive abilities or energy due to their medical conditions and treatment. The EPAT-16 can be utilised in routine care for continuous feedback on the overall degree of PC or for quality improvement. It can be used in large-scale multivariate studies assessing the relationship of a large number of constructs and their relationship to PC. It also enables large-scale collection of patient feedback across healthcare institutions, facilitating comparisons and reporting of PC levels.

Although other short questionnaires assessing PC exist,^36 37^ EPAT stands out due to its rigorous item development and selection process, which ensures a strong theoretical foundation and a comprehensive evaluation of PC,^38^ even with a reduced number of items.

### Limitations and future research

The results presented in this study are limited by the fact that all psychometric analyses were conducted in the same sample as had been used for item selection.^12^ It is also the same sample which we used to develop and evaluate the long version, EPAT-64. To confirm that the psychometric properties hold true when participants respond to only the 16 items, the EPAT-16 should be administered to a new, independent sample in future studies. Future research should specifically ask patients to complete both the EPAT-16 and EPAT-64 separately, allowing for an accurate calculation of the correlation between the two questionnaires.^12^ We did not report this correlation here, as using the same sample would have resulted in overestimation due to using responses on the EPAT-16 items on both sides of the calculation.^12^

Although the items were selected based on both content and psychometric characteristics - a key strength of the EPAT-16 - this process could have been more transparent by incorporating formal content rating and more representative by involving a larger group of experts in the selection process.^12^

Furthermore, most model fit indices were slightly outside the recommended range, suggesting that exploring alternatives to the unidimensional model may be worthwhile. The residual correlations and modification indices presented in Appendix 4, in conjunction with theoretical considerations, can guide the development of such alternative models. Wording of the EPAT-16 items was designed to be applicable for all patients regardless of their medical condition. Yet, measurement invariance between medical conditions was not supported in the data analysed here. Hence, while it can be applied generically to different healthcare institutions, comparability of results between different medical conditions is limited. Appendix 5 provides results of EPAT-16 sum scores per setting and medical condition to allow users to compare results of similar samples. Future research should assess the psychometric properties of the EPAT-16, including its factor structure and how it changes, in patient groups with further medical conditions that were not included here. In addition, the strong association of the sum score with the results from the Client Satisfaction Questionnaire deserves further attention, as the construct measured by the EPAT is likely to be a complex mixture of subjective and objective experiences.^35^ Finally, validating the English version of the EPAT-16, along with translating and testing it in other languages, would expand access to more patients and facilitate research on PC across different cultures, countries, and healthcare systems.

## Conclusions

The EPAT-16 provides a brief tool for assessing PC while covering all 16 dimensions from the integrative model of PC. Its concise format makes it well-suited for routine care settings, where time, resources, and patient capacity may be limited. It can be used for quality improvement, studies examining relationships of a large number of variates, and public reporting of PC across different healthcare institutions. Future research should focus on validating the EPAT-16 in independent samples, exploring its factor structure across various patient populations, and expanding its use across different languages and healthcare systems.

## Supporting information

Appendix 1 COSMIN reporting guideline

Appendix 2 Sample flow chart and characteristics

Appendix 3 Item characteristics per medical condition

Appendix 4 Residual correlations and modification indices

Appendix 5 Descriptive statistics of EPAT-16 sum score per group

## Data Availability

All data produced in the present study are available upon reasonable request to the authors.

## AUTHORS CONTRIBUTIONS

IS was the responsible principle investigator of the study. IS, LK and MH were involved in planning and preparation of the study. SZ, AS, HF and EC recruited participants and collected data. EC analysed data with supervision by LK. EC, SZ, JZ, IS, MH, PH and LK interpreted results and discussed the item selection. EC wrote the first draft of the manuscript. All authors critically revised the manuscript for important intellectual content. All authors gave final approval of the version to be published and agreed to be accountable for the work.

During the writing of the manuscript, EC used ChatGPT (OpenAI, Version 2) for language editing to improve readability and conciseness, since she is not a native English speaker. All content was created by the authors and only prompts to improve the language without altering the content were used. After using ChatGPT, all authors reviewed and edited the content as needed and takes full responsibility for the content of the publication.

## ETHICS APPROVAL AND CONSENT TO PARTICIPATE

The study was carried out according to the latest version of the Helsinki Declaration of the World Medical Association. Principles of good scientific practice were respected. The study had been approved by the Ethics Committee of the Medical Association Hamburg (study ID: PV5724). Study participation was voluntary and no foreseeable risks for participants resulted from the participation in this study. Participants were fully informed about the aims of the study, data collection, and the use of collected data. Written informed consent was obtained prior to participation. Preserving principles of data sensitivity, data protection, and confidentiality requirements were met.

## DATA AVAILABILITY

Data are available upon reasonable request.

