## Appendix 1 COSMIN reporting guideline for "Development and psychometric testing of EPAT-16: A short and valid measure for patient-centeredness from the patient’s perspective"

**Appendix 2:** Sample flow chart and characteristics

**Article:** Development and psychometric testing of EPAT-16: A short and valid measure for patient-centeredness from the patient's perspective

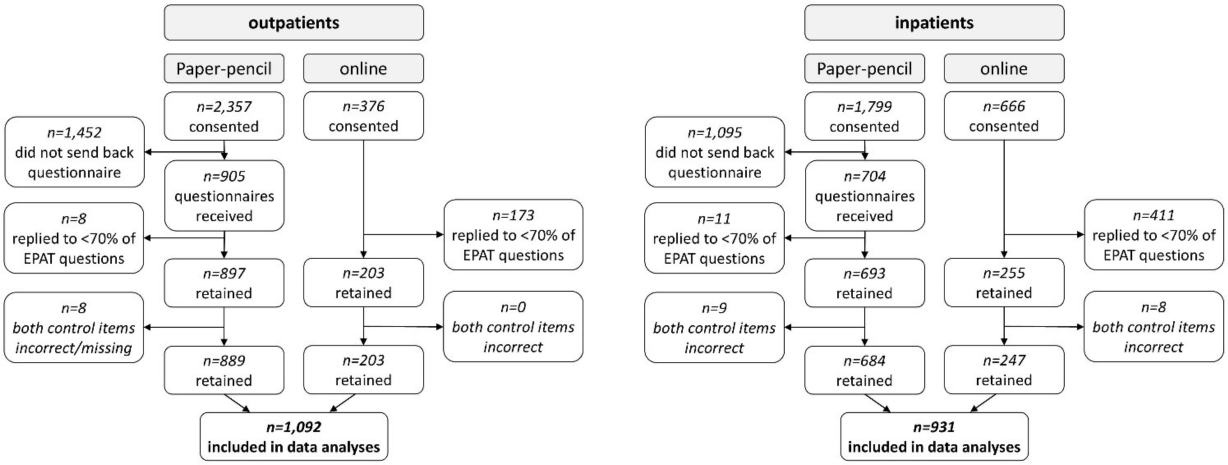


Figure 1: Sample flow chart (Originally published in Christalle et al., 2024[1], licensed under Creative Commons Attribution Non Commercial (CC BY-NC 4.0))

Table 1: Sample characteristics (Originally published in Christalle et al., 2024[1], licensed under Creative Commons Attribution Non Commercial (CC BY-NC 4.0))

| Characteristics | Outpatients | Inpatients |
|---|---|---|
| Total sample size | n = 1092 | n = 931 |
| In treatment for | | |
| Cardiovascular disease | n = 277 (25.4%) | n = 286 (30.7 %) |
| Cancer | n = 273 (25.0%) | n = 351 (37.7 %) |
| Musculoskeletal disease | n = 217 (19.9 %) | n = 92 (9.9 %) |
| Mental disorder | n = 273 (25.0 %) | n = 202 (21.7 %) |
| No information | n = 52 (4.8%)[+] | n = 0 (0 %) |
| Age (in years) | M = 53.1 (SD = 17.5) | M = 56.0 (SD = 17.6) |
| No response | n = 23 (2.1%) | n = 19 (2.0 %) |
| Years since initial diagnosis | M = 11.4 (SD = 12.0) | M = 8.5 (SD = 10.0) |
| No response | n = 124 (11.4%) | n = 118 (12.7 %) |
| Years as patient in this outpatient clinic | M = 4.53 (SD = 6.5) | - |
| No response | n = 129 (11.8%) | - |
| Length of stay (in days) | - | M = 18.1 (SD = 30.6) |
| No response | - | n = 74 (7.9 %) |
| Health literacy[a] | M = 50.7 (SD = 7.7) | M = 50.6 (SD = 7.7) |
| No response | n = 439 (40.2%) | n = 304 (32.7 %) |
| Satisfaction[b] | M = 27.5 (SD = 4.6) | M = 28.0 (SD = 4.9) |
| No response | n = 15 (1.4%) | n = 13 (1.4 %) |

| Characteristics | Outpatients | Inpatients |
|---|---|---|
| Health status[c] | M = 3.4 (SD = 2.8) | M = 3.4 (SD = 4.1) |
| No response | n = 44 (4.0%) | n = 36 (3.9 %) |
| Comorbidity (Do you have any further diseases?) | | |
| Yes | n = 551 (50.5 %) | n = 467 (50.2 %) |
| No | n = 450 (41.2 %) | n = 393 (42.2 %) |
| No response | n = 91 (8.3%) | n = 71 (7.6 %) |
| Gender | | |
| Female | n = 646 (59.2 %) | n = 384 (41.2 %) |
| Male | n = 420 (38.5 %) | n = 517 (55.5 %) |
| Diverse | n = 5 (0.5 %) | n = 6 (0.6 %) |
| No response | n = 21 (1.9%) | n = 24 (2.6 %) |
| Marital status | | |
| Unmarried and unpartnered | n = 343 (31.4 %) | n = 237 (25.5 %) |
| Married or partnered | n = 552 (50.5 %) | n = 524 (56.3 %) |
| Divorced | n = 107 (9.8 %) | n = 80 (8.6 %) |
| Widowed | n = 61 (5.6 %) | n = 54 (5.8 %) |
| No response | n = 29 (2.7%) | n = 36 (3.9 %) |
| Formal education | | |
| Low[d] | n = 10 (0.9 %) | n = 20 (2.1 %) |
| Intermediate[e] | n = 404 (37.0 %) | n = 383 (41.2 %) |
| High[f] | n = 267 (24.5 %) | n = 204 (21.9 %) |
| Very high[g] | n = 376 (34.4 %) | n = 279 (30.0 %) |
| No response | n = 23 (2.1%) | n = 27 (2.9 %) |
| Occupational status* | | |
| Employed | n = 447 (40.9 %) | n = 357 (38.3 %) |
| Unemployed | n = 66 (6.0 %) | n = 64 (6-9 %) |
| Student/trainee | n = 91 (8.3 %) | n = 50 (5.4 %) |
| Parental leave/stay at home | n = 57 (5.2 %) | n = 35 (3.8 %) |
| Retired | n = 394 (36.1 %) | n = 392 (42.1 %) |
| No response | n = 26 (2.4%) | n = 17 (1.8 %) |
| Health insurance* | | |
| Statutory | n = 910 (83.3 %) | n = 721 (77.4 %) |
| Private | n = 164 (15.0 %) | n = 214 (23.0 %) |
| No response | n = 32 (2.9%) | n = 34 (3.7 %) |
| Migration background (Were you or your parents born in another country than Germany?) | | |
| Yes | n = 167 (15.3 %) | n = 150 (16.1 %) |
| No | N = 898 (82.2 %) | n = 758 (81.4 %) |
| No response | n = 27 (2.5%) | n = 23 (2.5 %) |

Note: * multiple answers possible, M = Mean, SD = Standard Deviation, [+] These patients were recruited in a general practice. They assured us, that they were treated for one of the four health conditions, but failed to indicate which one in the questionnaire. [a] Health literacy measured by the total sum score of the HLS-EU-Q16[2], range 16-64, high value = high health literacy, [b] Satisfaction with care measured by ZUF-8[3], range 8-32, high value = high satisfaction, [b] General health status measured by first item of SF-12[4], range 0-5, high value = low health status, [d] low = no formal degree or graduation after less than 10years at school; [e] intermediate = graduation after 9 or 10years at school; [f] high = graduation after more than 10years at school; [g] very high = college or university degree
