## Appendix 2 Sample flow chart and characteristics for "Development and psychometric testing of EPAT-16: A short and valid measure for patient-centeredness from the patient’s perspective"

**Appendix 3: Item characteristics per medical condition**

**Article:** Development and psychometric testing of EPAT-16: A short and valid measure for patient-centeredness from the patient's perspective

**Abbreviations:**

- Card = Cardiovascular diseases
- Mus = Muscoloskeletal diseases
- Ment = Mental disorders

Table 1: Mean and standard deviation per medical condition – outpatient sample

| | | mean | | | | | standard deviation | | | | |
|---|---|---|---|---|---|---|---|---|---|---|---|
| | Item | **All** | Card | Cancer | Mus | Ment | **All** | Card | Cancer | Mus | Ment |
| Item 1 | The healthcare professionals were sensitive (for example they addressed my feelings, showed understanding, or empathized with my situation). | **5.1** | 5.1 | 5.1 | 4.8 | 5.1 | **1.2** | 1.2 | 1.2 | 1.5 | 1.2 |
| Item 2 | I trusted my healthcare professionals. | **5.2** | 5.3 | 5.3 | 5.0 | 5.3 | **1.1** | 1.1 | 1.0 | 1.3 | 1.0 |
| Item 3 | My wishes, needs and expectations were asked and taken into account in the treatment. | **4.8** | 4.7 | 4.7 | 4.5 | 4.7 | **1.4** | 1.4 | 1.4 | 1.6 | 1.4 |
| Item 4 | My entire personal life was taken into account during the treatment (for example, job, family and friends, partnership and sexuality, culture and religion, age, or financial circumstances). | **3.9** | 3.2 | 3.5 | 3.4 | 3.5 | **1.8** | 1.8 | 1.8 | 1.8 | 1.8 |
| Item 5 | I was given enough time to describe my concerns and my situation (for example, medical history or current symptoms). | **5.3** | 5.3 | 5.4 | 5.1 | 5.4 | **1.1** | 1.2 | 0.9 | 1.3 | 0.9 |
| Item 6 | I was asked if I use or would like to use additional services (for example, support groups, counseling, health courses, complementary and alternative medicine, or spiritual support/pastoral care). | **3.1** | 2.3 | 3.0 | 2.6 | 3.0 | **2.0** | 1.9 | 1.9 | 1.8 | 1.9 |
| Item 7 | The processes within the team were well organized. | **5.1** | 5.0 | 5.2 | 5.2 | 5.2 | **1.0** | 1.1 | 1.0 | 0.9 | 1.0 |
| Item 8 | If I wanted to speak to a physician, they were easily accessible. | **4.7** | 4.6 | 5.0 | 4.3 | 5.0 | **1.4** | 1.4 | 1.0 | 1.6 | 1.0 |
| Item 9 | It was discussed with me whether follow-up appointments would be useful (for example, for aftercare or further treatment). | **5.1** | 5.2 | 5.2 | 4.8 | 5.2 | **1.4** | 1.3 | 1.2 | 1.6 | 1.2 |
| Item 10 | I was encouraged to speak up if I noticed inconsistencies in my treatment. | **3.9** | 3.7 | 4.1 | 3.5 | 4.1 | **1.8** | 1.9 | 1.8 | 1.9 | 1.8 |
| Item 11 | I received information about my condition from my healthcare professionals (for example, causes, symptoms, effects or course). | **4.6** | 4.6 | 4.8 | 4.3 | 4.8 | **1.6** | 1.6 | 1.4 | 1.6 | 1.4 |
| Item 12 | I was an equal partner with my healthcare professionals (for example, in making decisions or sharing information). | **5.0** | 5.0 | 5.0 | 4.8 | 5.0 | **1.2** | 1.1 | 1.1 | 1.5 | 1.1 |
| Item 13 | I was informed about the options for involving my family members in the treatment (for example, accompanying to appointments, participating in conversations, or assisting with medication intake). | **3.1** | 3.0 | 3.4 | 2.2 | 3.4 | **2.0** | 2.0 | 2.0 | 1.7 | 2.0 |
| Item 14 | I was encouraged to improve my health by changing my behavior (for example, through diet, exercise, reducing tobacco or alcohol). | **4.0** | 3.5 | 3.9 | 3.9 | 3.9 | **1.8** | 1.9 | 1.8 | 1.7 | 1.8 |
| Item 15 | When I had pain, I was helped quickly. | **4.8** | 5.0 | 5.3 | 4.5 | 5.3 | **1.4** | 1.4 | 0.9 | 1.4 | 0.9 |
| Item 16 | The healthcare professionals addressed my fears and concerns (for example, by showing understanding and providing encouragement). | **4.4** | 3.8 | 4.2 | 3.9 | 4.2 | **1.7** | 1.9 | 1.6 | 1.7 | 1.6 |
| | EPAT-16 sum score | **72.6** | 70.9 | 73.5 | 68.3 | 76.6 | **16.0** | 15.7 | 14.9 | 18.2 | 13.9 |

Table 2: Further item characteristics per medical condition – outpatient sample

| | Item difficulty | | | | | 'does not concern me' | | | | | no reply | | | | | item total correlation | | | | |
|---|---|---|---|---|---|---|---|---|---|---|---|---|---|---|---|---|---|---|---|---|
| | **All** | Card | Cancer | Mus | Ment | **All** | Card | Cancer | Mus | Ment | **All** | Card | Cancer | Mus | Ment | **All** | Card | Cancer | Mus | Ment |
| Item 1 | **82.1** | 81.5 | 82.2 | 75.2 | 82.2 | **4.2%** | 8.3% | 4.4% | 2.8% | 4.4% | **1.1%** | 1.4% | 1.8% | 0.5% | 1.8% | **.714** | .652 | .672 | .805 | .680 |
| Item 2 | **84.8** | 86.0 | 86.1 | 79.2 | 86.1 | **1.8%** | 4.7% | 0.4% | 0.9% | 0.4% | **1.2%** | 1.1% | 1.5% | 0.9% | 1.5% | **.694** | .623 | .666 | .778 | .680 |
| Item 3 | **75.1** | 74.5 | 73.3 | 69.5 | 73.3 | **8.3%** | 14.1% | 10.6% | 3.7% | 10.6% | **0.8%** | 1.1% | 0.7% | 1.4% | 0.7% | **.748** | .689 | .687 | .827 | .760 |
| Item 4 | **57.0** | 43.9 | 49.4 | 48.6 | 49.4 | **16.1%** | 28.9% | 18.3% | 12.4% | 18.3% | **1.2%** | 1.8% | 1.8% | 0.5% | 1.8% | **.646** | .609 | .680 | .689 | .637 |
| Item 5 | **86.3** | 85.9 | 87.9 | 81.1 | 87.9 | **2.6%** | 5.1% | 2.2% | 2.3% | 2.2% | **1.0%** | 2.5% | 1.1% | 0.5% | 1.1% | **.673** | .850 | .583 | .787 | .625 |
| Item 6 | **42.3** | 26.9 | 39.5 | 31.5 | 39.5 | **30.4%** | 54.5% | 23.8% | 29.0% | 23.8% | **1.1%** | 1.8% | 1.8% | 0.5% | 1.8% | **.493** | .490 | .548 | .532 | .314 |
| Item 7 | **82.8** | 80.8 | 83.9 | 83.6 | 83.9 | **7.1%** | 3.6% | 3.7% | 2.8% | 3.7% | **0.8%** | 1.8% | 0.7% | 0.5% | 0.7% | **.403** | .415 | .369 | .429 | .418 |
| Item 8 | **73.1** | 72.6 | 80.4 | 66.0 | 80.4 | **23.4%** | 30.7% | 12.8% | 33.6% | 12.8% | **1.3%** | 1.4% | 2.2% | 0.5% | 2.2% | **.496** | .500 | .539 | .524 | .446 |
| Item 9 | **81.7** | 83.5 | 84.6 | 76.4 | 84.6 | **13.1%** | 16.2% | 12.8% | 11.1% | 12.8% | **1.4%** | 0.7% | 3.3% | 0.0% | 3.3% | **.479** | .478 | .488 | .486 | .465 |
| Item 10 | **57.5** | 54.1 | 61.6 | 50.6 | 61.6 | **23.4%** | 31.0% | 18.3% | 18.4% | 18.3% | **0.8%** | 0.7% | 1.5% | 0.9% | 1.5% | **.614** | .523 | .645 | .695 | .569 |
| Item 11 | **71.1** | 71.1 | 75.6 | 66.3 | 75.6 | **10.9%** | 15.5% | 10.3% | 6.5% | 10.3% | **0.7%** | 1.4% | 0.4% | 0.9% | 0.4% | **.630** | .643 | .555 | .729 | .645 |
| Item 12 | **79.4** | 80.2 | 79.5 | 76.1 | 79.5 | **7.6%** | 13.7% | 5.1% | 6.0% | 5.1% | **1.3%** | 1.4% | 1.1% | 1.8% | 1.1% | **.665** | .658 | .590 | .801 | .612 |
| Item 13 | **41.5** | 40.4 | 49.0 | 24.4 | 49.0 | **42.6%** | 55.2% | 30.8% | 49.8% | 30.8% | **1.3%** | 1.8% | 2.2% | 0.5% | 2.2% | **.553** | .536 | .583 | .581 | .502 |
| Item 14 | **60.9** | 50.7 | 58.6 | 58.9 | 58.6 | **26.0%** | 40.1% | 27.1% | 18.9% | 27.1% | **0.8%** | 1.1% | 1.1% | 0.9% | 1.1% | **.592** | .611 | .639 | .607 | .556 |
| Item 15 | **75.8** | 79.5 | 86.0 | 69.1 | 86.0 | **54.8%** | 69.7% | 52.7% | 19.8% | 52.7% | **1.0%** | 1.1% | 1.8% | 0.9% | 1.8% | **.551** | .476 | .529 | .668 | .572 |
| Item 16 | **67.8** | 56.9 | 63.2 | 58.6 | 63.2 | **19.2%** | 36.1% | 18.7% | 19.4% | 18.7% | **0.8%** | 0.7% | 1.5% | 0.5% | 1.5% | **.731** | .688 | .721 | .788 | .744 |

Notes: item total correlation = Corrected item total correlation with the other EPAT-16 items

Table 3: Mean and standard deviation per medical condition – inpatient sample

| | | mean | | | | | standard deviation | | | | |
|---|---|---|---|---|---|---|---|---|---|---|---|
| | Item | **All** | Card | Cancer | Mus | Ment | **All** | Card | Cancer | Mus | Ment |
| Item 1 | The healthcare professionals were sensitive (for example they addressed my feelings, showed understanding, or empathized with my situation). | **5.1** | 5.1 | 5.5 | 4.6 | 5.5 | **1.2** | 1.2 | 0.8 | 1.3 | 0.8 |
| Item 2 | I trusted my healthcare professionals. | **5.3** | 5.4 | 5.6 | 4.9 | 5.6 | **1.1** | 1.0 | 0.8 | 1.3 | 0.8 |
| Item 3 | My wishes, needs and expectations were asked and taken into account in the treatment. | **4.8** | 4.8 | 5.1 | 4.3 | 5.1 | **1.3** | 1.4 | 1.2 | 1.6 | 1.2 |
| Item 4 | My entire personal life was taken into account during the treatment (for example, job, family and friends, partnership and sexuality, culture and religion, age, or financial circumstances). | **3.8** | 3.3 | 3.8 | 3.5 | 3.8 | **1.8** | 1.7 | 1.8 | 1.9 | 1.8 |
| Item 5 | I was given enough time to describe my concerns and my situation (for example, medical history or current symptoms). | **5.2** | 5.3 | 5.5 | 4.8 | 5.5 | **1.1** | 1.0 | 0.9 | 1.3 | 0.9 |
| Item 6 | I was asked if I use or would like to use additional services (for example, support groups, counseling, health courses, complementary and alternative medicine, or spiritual support/pastoral care). | **3.4** | 2.5 | 3.8 | 2.8 | 3.8 | **2.0** | 1.8 | 2.0 | 1.8 | 2.0 |
| Item 7 | The processes within the team were well organized. | **5.1** | 5.2 | 5.4 | 4.9 | 5.4 | **1.1** | 1.1 | 0.9 | 1.2 | 0.9 |
| Item 8 | If I wanted to speak to a physician, they were easily accessible. | **4.8** | 4.9 | 5.2 | 4.5 | 5.2 | **1.2** | 1.2 | 1.0 | 1.4 | 1.0 |
| Item 9 | It was discussed with me whether follow-up appointments would be useful (for example, for aftercare or further treatment). | **5.0** | 5.0 | 5.3 | 4.8 | 5.3 | **1.4** | 1.4 | 1.2 | 1.5 | 1.2 |
| Item 10 | I was encouraged to speak up if I noticed inconsistencies in my treatment. | **4.1** | 4.1 | 4.3 | 3.7 | 4.3 | **1.7** | 1.7 | 1.8 | 1.8 | 1.8 |
| Item 11 | I received information about my condition from my healthcare professionals (for example, causes, symptoms, effects or course). | **4.6** | 4.8 | 4.7 | 4.4 | 4.7 | **1.5** | 1.4 | 1.4 | 1.7 | 1.4 |
| Item 12 | I was an equal partner with my healthcare professionals (for example, in making decisions or sharing information). | **4.8** | 4.8 | 5.2 | 4.4 | 5.2 | **1.3** | 1.3 | 1.0 | 1.5 | 1.0 |
| Item 13 | I was informed about the options for involving my family members in the treatment (for example, accompanying to appointments, participating in conversations, or assisting with medication intake). | **3.3** | 3.3 | 3.4 | 2.7 | 3.4 | **1.9** | 2.0 | 1.9 | 1.8 | 1.9 |
| Item 14 | I was encouraged to improve my health by changing my behavior (for example, through diet, exercise, reducing tobacco or alcohol). | **4.0** | 3.8 | 3.8 | 3.6 | 3.8 | **1.7** | 1.8 | 1.8 | 1.7 | 1.8 |
| Item 15 | When I had pain, I was helped quickly. | **5.4** | 5.6 | 5.7 | 4.9 | 5.7 | **1.0** | 0.8 | 0.7 | 1.3 | 0.7 |
| Item 16 | The healthcare professionals addressed my fears and concerns (for example, by showing understanding and providing encouragement). | **4.4** | 4.1 | 4.7 | 3.6 | 4.7 | **1.6** | 1.7 | 1.4 | 1.8 | 1.4 |
| | EPAT-16 sum score | **73.6** | 72.8 | 77.5 | 66.9 | 71.0 | **15.3** | 14.3 | 13.5 | 18.4 | 16.6 |

Table 4: Further item characteristics per medical condition - inpatient sample

| | **Item difficulty** | | | | | **'does not concern me'** | | | | | **no reply** | | | | | **item total correlation** | | | | |
|---|---|---|---|---|---|---|---|---|---|---|---|---|---|---|---|---|---|---|---|---|
| | **All** | Card | Cancer | Mus | Ment | **All** | Card | Cancer | Mus | Ment | **All** | Card | Cancer | Mus | Ment | **All** | Card | Cancer | Mus | Ment |
| Item 1 | **81.9** | 82.5 | 89.0 | 71.6 | 89.0 | **3.9%** | 7.7% | 2.3% | 5.4% | 2.3% | **0.8%** | 0.7% | 0.9% | 1.1% | 0.9% | **.701** | .585 | .649 | .791 | .797 |
| Item 2 | **86.3** | 88.6 | 91.2 | 78.7 | 91.2 | **1.2%** | 2.4% | 0.3% | 2.2% | 0.3% | **0.4%** | 1.0% | 0.3% | 0.0% | 0.3% | **.676** | .537 | .639 | .804 | .758 |
| Item 3 | **75.8** | 76.9 | 81.5 | 65.2 | 81.5 | **5.5%** | 9.4% | 5.9% | 3.3% | 5.9% | **0.9%** | 1.0% | 0.6% | 0.0% | 0.6% | **.742** | .646 | .695 | .845 | .817 |
| Item 4 | **55.8** | 45.8 | 55.3 | 49.1 | 55.3 | **14.2%** | 24.1% | 15.8% | 4.3% | 15.8% | **0.8%** | 0.3% | 0.9% | 2.2% | 0.9% | **.593** | .615 | .660 | .620 | .715 |
| Item 5 | **85.0** | 87.0 | 89.9 | 76.5 | 89.9 | **2.8%** | 2.1% | 5.3% | 0.0% | 5.3% | **0.5%** | 0.3% | 0.9% | 0.0% | 0.9% | **.690** | .616 | .645 | .750 | .750 |
| Item 6 | **48.7** | 30.0 | 56.8 | 35.5 | 56.8 | **25.8%** | 43.7% | 21.4% | 21.7% | 21.4% | **1.5%** | 1.0% | 2.3% | 1.1% | 2.3% | **.531** | .552 | .550 | .597 | .538 |
| Item 7 | **81.8** | 83.9 | 88.6 | 78.0 | 88.6 | **1.2%** | 1.4% | 0.3% | 3.3% | 0.3% | **1.0%** | 1.4% | 0.6% | 1.1% | 0.6% | **.605** | .557 | .625 | .600 | .645 |
| Item 8 | **76.2** | 78.0 | 83.5 | 69.5 | 83.5 | **6.6%** | 9.8% | 5.6% | 6.5% | 5.6% | **1.0%** | 1.0% | 1.5% | 0.0% | 1.5% | **.611** | .539 | .605 | .708 | .607 |
| Item 9 | **80.0** | 79.9 | 86.3 | 75.1 | 86.3 | **7.2%** | 8.4% | 7.3% | 2.2% | 7.3% | **1.0%** | 1.4% | 1.2% | 1.1% | 1.2% | **.547** | .470 | .463 | .619 | .650 |
| Item 10 | **61.9** | 62.0 | 65.5 | 53.5 | 65.5 | **14.2%** | 15.0% | 19.1% | 8.7% | 19.1% | **1.2%** | 1.0% | 1.5% | 1.1% | 1.5% | **.610** | .592 | .551 | .745 | .632 |
| Item 11 | **72.1** | 76.6 | 73.7 | 68.1 | 73.7 | **6.4%** | 5.2% | 7.9% | 5.4% | 7.9% | **1.3%** | 1.4% | 1.8% | 1.1% | 1.8% | **.606** | .578 | .606 | .719 | .627 |
| Item 12 | **75.8** | 75.1 | 83.6 | 68.5 | 83.6 | **4.4%** | 7.3% | 3.8% | 2.2% | 3.8% | **1.8%** | 2.1% | 2.6% | 1.1% | 2.6% | **.643** | .581 | .560 | .695 | .715 |
| Item 13 | **46.0** | 45.4 | 48.7 | 33.0 | 48.7 | **29.6%** | 39.2% | 30.5% | 28.3% | 30.5% | **1.0%** | 1.0% | 1.2% | 0.0% | 1.2% | **.578** | .590 | .618 | .662 | .471 |
| Item 14 | **60.8** | 56.0 | 56.5 | 52.6 | 56.5 | **25.8%** | 29.0% | 34.0% | 20.7% | 34.0% | **0.5%** | 0.3% | 1.2% | 0.0% | 1.2% | **.492** | .484 | .577 | .565 | .537 |
| Item 15 | **87.1** | 91.3 | 94.0 | 77.0 | 94.0 | **19.4%** | 23.8% | 17.0% | 5.4% | 17.0% | **0.8%** | 1.4% | 0.6% | 0.0% | 0.6% | **.492** | .455 | .440 | .561 | .505 |
| Item 16 | **68.8** | 62.3 | 74.5 | 51.3 | 74.5 | **15.9%** | 25.2% | 17.9% | 14.1% | 17.9% | **1.5%** | 0.7% | 2.6% | 1.1% | 2.6% | **.688** | .633 | .660 | .755 | .815 |

Notes: item total correlation = Corrected item total correlation with the other EPAT-16 items
