## Appendix 3 Item characteristics per medical condition for "Development and psychometric testing of EPAT-16: A short and valid measure for patient-centeredness from the patient’s perspective"

**Appendix 4: Residual correlations and modification indices for the unidimensional model**

**Article:** Development and psychometric testing of EPAT-16: A short and valid measure for patient-centeredness from the patient's perspective

Item abbreviations used in tables below:

| Abbreviation | Dimension | Item |
|---|---|---|
| Essential char | Essential char of the clinician | The healthcare professionals were sensitive (for example they addressed my feelings, showed understanding, or empathized with my situation). |
| Relationship | Clinician-patient relationship | I trusted my healthcare professionals. |
| Unique person | Patient as a unique person | My wishes, needs and expectations were asked and taken into account in the treatment. |
| Biopsychosocial | Biopsychosocial perspective | My entire personal life was taken into account during the treatment (for example, job, family and friends, partnership and sexuality, culture and religion, age, or financial circumstances). |
| Communication | Clinician-patient communication | I was given enough time to describe my concerns and my situation (for example, medical history or current symptoms). |
| Integration | Integration of medical and non-medical care | I was asked if I use or would like to use additional services (for example, support groups, counseling, health courses, complementary and alternative medicine, or spiritual support/pastoral care). |
| Teamwork | Teamwork and teambuilding | The processes within the team were well organized. |
| Access | Access to care | If I wanted to speak to a physician, they were easily accessible. |
| Coordination | Coordination and continuity of care | It was discussed with me whether follow-up appointments would be useful (for example, for aftercare or further treatment). |
| Safety | Patient safety | I was encouraged to speak up if I noticed inconsistencies in my treatment. |
| Information | Patient information | I received information about my condition from my healthcare professionals (for example, causes, symptoms, effects or course). |
| Involvement | Patient involvement in care | I was an equal partner with my healthcare professionals (for example, in making decisions or sharing information). |
| Family | Involvement of family and friends | I was informed about the options for involving my family members in the treatment (for example, accompanying to appointments, participating in conversations, or assisting with medication intake). |
| Empowerment | Patient empowerment | I was encouraged to improve my health by changing my behavior (for example, through diet, exercise, reducing tobacco or alcohol). |
| Physical | Physical support | When I had pain, I was helped quickly. |
| Emotional | Emotional support | The healthcare professionals addressed my fears and concerns (for example, by showing understanding and providing encouragement). |

Table 1: Residual correlations in the unidimensional model – outpatient sample

| | Essential char | Relation-ship | Unique person | Biopsycho-social | Communi-cation | Integration | Teamwork | Access | Coordi-nation | Safety | Information | Involvement | Family | Empower-ment | Physical | Emotional |
|---|---|---|---|---|---|---|---|---|---|---|---|---|---|---|---|---|
| Essential char | 0 | 0.059 | -0.005 | -0.043 | 0.021 | -0.063 | -0.022 | -0.001 | -0.047 | -0.042 | -0.028 | 0.01 | -0.092 | -0.084 | 0.01 | 0.04 |
| Relationship | 0.059 | 0 | -0.001 | -0.075 | 0.034 | -0.068 | -0.011 | -0.017 | 0.01 | -0.061 | -0.025 | 0.044 | -0.051 | -0.095 | 0.088 | -0.034 |
| Unique person | -0.005 | -0.001 | 0 | -0.011 | 0.021 | 0.013 | -0.052 | -0.04 | -0.021 | 0.031 | 0.013 | 0.037 | -0.023 | -0.026 | -0.041 | -0.009 |
| Biopsychosocial | -0.043 | -0.075 | -0.011 | 0 | -0.077 | 0.219 | -0.024 | -0.031 | -0.007 | 0.122 | 0.013 | -0.031 | 0.194 | 0.157 | -0.049 | 0.141 |
| Communication | 0.021 | 0.034 | 0.021 | -0.077 | 0 | -0.057 | 0.03 | 0.01 | 0.007 | -0.03 | -0.009 | 0.046 | -0.095 | -0.051 | 0.022 | -0.057 |
| Integration | -0.063 | -0.068 | 0.013 | 0.219 | -0.057 | 0 | 0 | -0.001 | 0.01 | 0.133 | 0.009 | -0.065 | 0.249 | 0.213 | 0.003 | 0.081 |
| Teamwork | -0.022 | -0.011 | -0.052 | -0.024 | 0.03 | 0 | 0 | 0.161 | 0.092 | 0.038 | 0.032 | -0.017 | 0.031 | 0.055 | 0.048 | -0.045 |
| Access | -0.001 | -0.017 | -0.04 | -0.031 | 0.01 | -0.001 | 0.161 | 0 | -0.022 | 0.001 | 0.009 | 0.033 | 0.066 | 0.007 | 0.058 | -0.038 |
| Coordination | -0.047 | 0.01 | -0.021 | -0.007 | 0.007 | 0.01 | 0.092 | -0.022 | 0 | 0.018 | 0.126 | -0.008 | 0.023 | 0.035 | -0.019 | -0.017 |
| Safety | -0.042 | -0.061 | 0.031 | 0.122 | -0.03 | 0.133 | 0.038 | 0.001 | 0.018 | 0 | 0.044 | -0.049 | 0.184 | 0.117 | 0.007 | 0.009 |
| Information | -0.028 | -0.025 | 0.013 | 0.013 | -0.009 | 0.009 | 0.032 | 0.009 | 0.126 | 0.044 | 0 | -0.051 | 0.125 | 0.123 | -0.042 | -0.005 |
| Involvement | 0.01 | 0.044 | 0.037 | -0.031 | 0.046 | -0.065 | -0.017 | 0.033 | -0.008 | -0.049 | -0.051 | 0 | -0.115 | -0.059 | 0 | -0.037 |
| Family | -0.092 | -0.051 | -0.023 | 0.194 | -0.095 | 0.249 | 0.031 | 0.066 | 0.023 | 0.184 | 0.125 | -0.115 | 0 | 0.214 | 0.062 | 0.067 |
| Empowerment | -0.084 | -0.095 | -0.026 | 0.157 | -0.051 | 0.213 | 0.055 | 0.007 | 0.035 | 0.117 | 0.123 | -0.059 | 0.214 | 0 | -0.055 | 0.074 |
| Physical | 0.01 | 0.088 | -0.041 | -0.049 | 0.022 | 0.003 | 0.048 | 0.058 | -0.019 | 0.007 | -0.042 | 0 | 0.062 | -0.055 | 0 | -0.035 |
| Emotional | 0.04 | -0.034 | -0.009 | 0.141 | -0.057 | 0.081 | -0.045 | -0.038 | -0.017 | 0.009 | -0.005 | -0.037 | 0.067 | 0.074 | -0.035 | 0 |

Table 2: Residual correlations in the unidimensional model – inpatient sample

| | Essential char | Relation-ship | Unique person | Biopsycho-social | Communi-cation | Integration | Teamwork | Access | Coordi-nation | Safety | Information | Involvement | Family | Empower-ment | Physical | Emotional |
|---|---|---|---|---|---|---|---|---|---|---|---|---|---|---|---|---|
| Essential char | 0 | 0.034 | -0.02 | -0.062 | 0.023 | -0.035 | 0.045 | 0.04 | -0.005 | -0.035 | -0.067 | 0.004 | -0.074 | -0.068 | 0.026 | 0.02 |
| Relationship | 0.034 | 0 | 0.001 | -0.078 | 0.035 | -0.089 | 0.059 | -0.021 | 0.016 | -0.077 | -0.021 | 0.05 | -0.107 | -0.061 | 0.021 | -0.018 |
| Unique person | -0.02 | 0.001 | 0 | 0.019 | 0.003 | 0.014 | -0.02 | 0.012 | -0.018 | 0.023 | 0.007 | 0.025 | 0.027 | 0.027 | 0.007 | -0.048 |
| Biopsychosocial | -0.062 | -0.078 | 0.019 | 0 | -0.047 | 0.247 | -0.093 | -0.06 | -0.012 | 0.121 | 0.098 | -0.039 | 0.217 | 0.263 | -0.127 | 0.204 |
| Communication | 0.023 | 0.035 | 0.003 | -0.047 | 0 | -0.056 | 0.005 | -0.011 | 0.014 | -0.026 | 0.017 | -0.001 | -0.06 | -0.047 | 0.032 | -0.032 |
| Integration | -0.035 | -0.089 | 0.014 | 0.247 | -0.056 | 0 | -0.069 | -0.05 | 0.061 | 0.167 | 0.031 | -0.052 | 0.278 | 0.22 | -0.065 | 0.167 |
| Teamwork | 0.045 | 0.059 | -0.02 | -0.093 | 0.005 | -0.069 | 0 | 0.07 | 0.015 | -0.031 | -0.024 | -0.025 | -0.034 | -0.072 | 0.046 | -0.071 |
| Access | 0.04 | -0.021 | 0.012 | -0.06 | -0.011 | -0.05 | 0.07 | 0 | 0.031 | -0.036 | -0.05 | 0.002 | -0.038 | -0.05 | 0.075 | -0.019 |
| Coordination | -0.005 | 0.016 | -0.018 | -0.012 | 0.014 | 0.061 | 0.015 | 0.031 | 0 | -0.039 | 0.045 | -0.03 | 0.043 | -0.031 | -0.036 | 0 |
| Safety | -0.035 | -0.077 | 0.023 | 0.121 | -0.026 | 0.167 | -0.031 | -0.036 | -0.039 | 0 | 0.07 | -0.013 | 0.23 | 0.159 | -0.057 | 0.035 |
| Information | -0.067 | -0.021 | 0.007 | 0.098 | 0.017 | 0.031 | -0.024 | -0.05 | 0.045 | 0.07 | 0 | 0.006 | 0.127 | 0.066 | -0.045 | 0.004 |
| Involvement | 0.004 | 0.05 | 0.025 | -0.039 | -0.001 | -0.052 | -0.025 | 0.002 | -0.03 | -0.013 | 0.006 | 0 | -0.041 | -0.061 | 0.011 | -0.006 |
| Family | -0.074 | -0.107 | 0.027 | 0.217 | -0.06 | 0.278 | -0.034 | -0.038 | 0.043 | 0.23 | 0.127 | -0.041 | 0 | 0.218 | -0.059 | 0.107 |
| Empowerment | -0.068 | -0.061 | 0.027 | 0.263 | -0.047 | 0.22 | -0.072 | -0.05 | -0.031 | 0.159 | 0.066 | -0.061 | 0.218 | 0 | -0.1 | 0.1 |
| Physical | 0.026 | 0.021 | 0.007 | -0.127 | 0.032 | -0.065 | 0.046 | 0.075 | -0.036 | -0.057 | -0.045 | 0.011 | -0.059 | -0.1 | 0 | -0.061 |
| Emotional | 0.02 | -0.018 | -0.048 | 0.204 | -0.032 | 0.167 | -0.071 | -0.019 | 0 | 0.035 | 0.004 | -0.006 | 0.107 | 0.1 | -0.061 | 0 |

Table 3: Modification indices in the unidimensional model – outpatient sample

| lhs | op | rhs | mi | epc | sepc.lv | sepc.all | sepc.nox |
|---|---|---|---|---|---|---|---|
| Biopsychosocial | ~~ | Integration | 57.9 | 0.745 | 0.745 | 0.300 | 0.300 |
| Biopsychosocial | ~~ | Emotional | 55.3 | 0.426 | 0.426 | 0.288 | 0.288 |
| Integration | ~~ | Family | 47.4 | 0.915 | 0.915 | 0.308 | 0.308 |
| Integration | ~~ | Empowerment | 45.9 | 0.712 | 0.712 | 0.279 | 0.279 |
| Essential char | ~~ | Relationship | 44.7 | 0.124 | 0.124 | 0.257 | 0.257 |
| Biopsychosocial | ~~ | Family | 42.3 | 0.660 | 0.660 | 0.281 | 0.281 |
| Biopsychosocial | ~~ | Empowerment | 38.1 | 0.486 | 0.486 | 0.241 | 0.241 |
| Family | ~~ | Empowerment | 37.8 | 0.671 | 0.671 | 0.278 | 0.278 |
| Teamwork | ~~ | Access | 34.8 | 0.236 | 0.236 | 0.218 | 0.218 |
| Coordination | ~~ | Information | 33.6 | 0.288 | 0.288 | 0.207 | 0.207 |
| Safety | ~~ | Family | 30.6 | 0.609 | 0.609 | 0.250 | 0.250 |
| Information | ~~ | Empowerment | 28.7 | 0.347 | 0.347 | 0.205 | 0.205 |
| Relationship | ~~ | Biopsychosocial | 27.6 | -0.188 | -0.188 | -0.195 | -0.195 |
| Relationship | ~~ | Empowerment | 27.3 | -0.202 | -0.202 | -0.205 | -0.205 |
| Involvement | ~~ | Family | 27.0 | -0.309 | -0.309 | -0.231 | -0.231 |
| Communication | ~~ | Family | 25.3 | -0.261 | -0.261 | -0.220 | -0.220 |
| Biopsychosocial | ~~ | Safety | 24.4 | 0.389 | 0.389 | 0.191 | 0.191 |
| Biopsychosocial | ~~ | Communication | 24.3 | -0.178 | -0.178 | -0.180 | -0.180 |
| Essential char | ~~ | Empowerment | 23.5 | -0.197 | -0.197 | -0.194 | -0.194 |
| Essential char | ~~ | Family | 23.4 | -0.258 | -0.258 | -0.218 | -0.218 |
| Essential char | ~~ | Emotional | 20.9 | 0.138 | 0.138 | 0.185 | 0.185 |
| Relationship | ~~ | Integration | 19.5 | -0.214 | -0.214 | -0.176 | -0.176 |
| Essential char | ~~ | Integration | 18.6 | -0.219 | -0.219 | -0.176 | -0.176 |
| Communication | ~~ | Emotional | 18.2 | -0.124 | -0.124 | -0.166 | -0.166 |
| Relationship | ~~ | Safety | 17.2 | -0.161 | -0.161 | -0.161 | -0.161 |
| Relationship | ~~ | Physical | 16.8 | 0.146 | 0.146 | 0.208 | 0.208 |
| Integration | ~~ | Safety | 16.2 | 0.427 | 0.427 | 0.166 | 0.166 |
| Safety | ~~ | Empowerment | 15.7 | 0.334 | 0.334 | 0.160 | 0.160 |
| Relationship | ~~ | Involvement | 14.7 | 0.079 | 0.079 | 0.143 | 0.143 |
| Communication | ~~ | Involvement | 14.7 | 0.079 | 0.079 | 0.141 | 0.141 |
| Unique person | ~~ | Involvement | 14.4 | 0.093 | 0.093 | 0.149 | 0.149 |
| Information | ~~ | Family | 14.0 | 0.317 | 0.317 | 0.161 | 0.161 |
| Integration | ~~ | Involvement | 13.2 | -0.207 | -0.207 | -0.146 | -0.146 |
| Teamwork | ~~ | Coordination | 12.8 | 0.137 | 0.137 | 0.124 | 0.124 |
| Information | ~~ | Involvement | 12.8 | -0.123 | -0.123 | -0.131 | -0.131 |
| Unique person | ~~ | Teamwork | 12.3 | -0.094 | -0.094 | -0.129 | -0.129 |
| Essential char | ~~ | Biopsychosocial | 12.1 | -0.130 | -0.130 | -0.132 | -0.132 |
| Communication | ~~ | Integration | 12.0 | -0.171 | -0.171 | -0.136 | -0.136 |
| Essential char | ~~ | Coordination | 10.9 | -0.102 | -0.102 | -0.123 | -0.123 |
| Safety | ~~ | Involvement | 10.6 | -0.147 | -0.147 | -0.127 | -0.127 |
| Involvement | ~~ | Empowerment | 10.2 | -0.145 | -0.145 | -0.126 | -0.126 |
| Relationship | ~~ | Family | 10.2 | -0.162 | -0.162 | -0.141 | -0.141 |

lhs = left hand side; rhs = right hand side; mi = modification indice; epc = expected parameter change; sepc.lv = only standardizing the latent variables; sepc.al = standardizing all variables; sepc.nox = standardizing all but exogenous observed variables

Table 4: Modification indices in the unidimensional model – inpatient sample

| lhs | op | rhs | mi | epc | sepc.lv | sepc.all | sepc.nox |
|---|---|---|---|---|---|---|---|
| Biopsychosocial | ~~ | Emotional | 75.6 | 0.574 | 0.574 | 0.345 | 0.345 |
| Integration | ~~ | Family | 73.3 | 1.056 | 1.056 | 0.375 | 0.375 |
| Biopsychosocial | ~~ | Empowerment | 68.3 | 0.769 | 0.769 | 0.342 | 0.342 |
| Biopsychosocial | ~~ | Integration | 66.2 | 0.849 | 0.849 | 0.334 | 0.334 |
| Biopsychosocial | ~~ | Family | 59.9 | 0.777 | 0.777 | 0.323 | 0.323 |
| Safety | ~~ | Family | 54.8 | 0.721 | 0.721 | 0.317 | 0.317 |
| Relationship | ~~ | Family | 44.2 | -0.310 | -0.310 | -0.284 | -0.284 |
| Integration | ~~ | Empowerment | 42.4 | 0.747 | 0.747 | 0.284 | 0.284 |
| Family | ~~ | Empowerment | 40.7 | 0.707 | 0.707 | 0.284 | 0.284 |
| Integration | ~~ | Emotional | 36.9 | 0.501 | 0.501 | 0.257 | 0.257 |
| Relationship | ~~ | Integration | 32.1 | -0.273 | -0.273 | -0.236 | -0.236 |
| Safety | ~~ | Empowerment | 29.8 | 0.485 | 0.485 | 0.228 | 0.228 |
| Essential char | ~~ | Family | 27.7 | -0.263 | -0.263 | -0.229 | -0.229 |
| Integration | ~~ | Safety | 27.4 | 0.529 | 0.529 | 0.220 | 0.220 |
| Biopsychosocial | ~~ | Physical | 26.4 | -0.253 | -0.253 | -0.209 | -0.209 |
| Information | ~~ | Family | 26.1 | 0.412 | 0.412 | 0.214 | 0.214 |
| Biopsychosocial | ~~ | Teamwork | 25.1 | -0.227 | -0.227 | -0.190 | -0.190 |
| Relationship | ~~ | Biopsychosocial | 23.5 | -0.186 | -0.186 | -0.188 | -0.188 |
| Essential char | ~~ | Information | 23.1 | -0.155 | -0.155 | -0.186 | -0.186 |
| Relationship | ~~ | Safety | 22.0 | -0.172 | -0.172 | -0.185 | -0.185 |
| Biopsychosocial | ~~ | Safety | 21.6 | 0.375 | 0.375 | 0.183 | 0.183 |
| Teamwork | ~~ | Access | 19.7 | 0.120 | 0.120 | 0.166 | 0.166 |
| Teamwork | ~~ | Emotional | 19.2 | -0.157 | -0.157 | -0.173 | -0.173 |
| Biopsychosocial | ~~ | Information | 18.9 | 0.290 | 0.290 | 0.166 | 0.166 |
| Relationship | ~~ | Teamwork | 18.1 | 0.087 | 0.087 | 0.160 | 0.160 |
| Family | ~~ | Emotional | 17.3 | 0.331 | 0.331 | 0.180 | 0.180 |
| Relationship | ~~ | Involvement | 15.9 | 0.092 | 0.092 | 0.154 | 0.154 |
| Essential char | ~~ | Biopsychosocial | 15.3 | -0.161 | -0.161 | -0.155 | -0.155 |
| Essential char | ~~ | Empowerment | 14.4 | -0.173 | -0.173 | -0.160 | -0.160 |
| Access | ~~ | Physical | 13.8 | 0.111 | 0.111 | 0.150 | 0.150 |
| Essential char | ~~ | Teamwork | 13.4 | 0.081 | 0.081 | 0.141 | 0.141 |
| Empowerment | ~~ | Physical | 13.0 | -0.197 | -0.197 | -0.157 | -0.157 |
| Integration | ~~ | Teamwork | 12.4 | -0.200 | -0.200 | -0.144 | -0.144 |
| Integration | ~~ | Involvement | 11.6 | -0.217 | -0.217 | -0.142 | -0.142 |
| Unique person | ~~ | Emotional | 11.4 | -0.125 | -0.125 | -0.140 | -0.140 |
| Empowerment | ~~ | Emotional | 10.9 | 0.243 | 0.243 | 0.141 | 0.141 |
| Teamwork | ~~ | Empowerment | 10.8 | -0.164 | -0.164 | -0.133 | -0.133 |
| Communication | ~~ | Family | 10.4 | -0.157 | -0.157 | -0.138 | -0.138 |
| Essential char | ~~ | Access | 10.3 | 0.080 | 0.080 | 0.127 | 0.127 |

lhs = left hand side; rhs = right hand side; mi = modification indice; epc = expected parameter change; sepc.lv = only standardizing the latent variables; sepc.al = standardizing all variables; sepc.nox = standardizing all but exogenous observed variables
