## Appendix 4 Residual correlations and modification indices for "Development and psychometric testing of EPAT-16: A short and valid measure for patient-centeredness from the patient’s perspective"

**Appendix 5: Descriptive statistics of EPAT-16 sum score per group**

**Article:** Development and psychometric testing of EPAT-16: A short and valid measure for patient-centeredness from the patient's perspective

| | Outpatient sample | | | | | Inpatient sample | | | | |
|---|---|---|---|---|---|---|---|---|---|---|
| | All | Card | Cancer | Mus | Ment | All | Card | Cancer | Mus | Ment |
| **M** | 72.6 | 70.9 | 73.5 | 68.3 | 76.6 | 73.6 | 72.8 | 77.5 | 66.9 | 71.0 |
| **SD** | 15.7 | 15.3 | 14.7 | 18.0 | 13.7 | 15.2 | 14.1 | 13.4 | 18.2 | 16.5 |
| **Percentiles** | | | | | | | | | | |
| 5 | 42 | 45 | 46 | 35 | 47 | 46 | 50 | 53 | 31 | 38 |
| 10 | 50 | 51 | 55 | 42 | 59 | 53 | 54 | 59 | 36 | 48 |
| 15 | 56 | 55 | 58 | 47 | 64 | 58 | 59 | 63 | 46 | 54 |
| 20 | 60 | 58 | 61 | 52 | 67 | 62 | 61 | 65 | 50 | 57 |
| 25 | 63 | 61 | 64 | 56 | 69 | 64 | 64 | 69 | 55 | 63 |
| 30 | 66 | 63 | 66 | 59 | 71 | 67 | 66 | 71 | 60 | 65 |
| 35 | 68 | 65 | 68 | 64 | 74 | 70 | 68 | 74 | 62 | 67 |
| 40 | 70 | 68 | 71 | 66 | 76 | 72 | 70 | 76 | 64 | 70 |
| 45 | 73 | 70 | 73 | 68 | 78 | 74 | 72 | 78 | 66 | 72 |
| 50 | 75 | 72 | 75 | 71 | 79 | 76 | 73 | 80 | 69 | 74 |
| 55 | 77 | 74 | 77 | 74 | 81 | 78 | 76 | 81 | 73 | 75 |
| 60 | 79 | 77 | 79 | 77 | 82 | 80 | 78 | 83 | 76 | 77 |
| 65 | 81 | 79 | 80 | 79 | 84 | 82 | 79 | 85 | 78 | 80 |
| 70 | 83 | 81 | 83 | 80 | 86 | 84 | 82 | 87 | 80 | 82 |
| 75 | 85 | 83 | 85 | 82 | 87 | 86 | 84 | 88 | 82 | 84 |
| 80 | 87 | 85 | 87 | 85 | 89 | 88 | 86 | 90 | 84 | 86 |
| 85 | 89 | 88 | 90 | 87 | 90 | 90 | 88 | 91 | 85 | 88 |
| 90 | 91 | 90 | 92 | 90 | 92 | 92 | 91 | 93 | 89 | 90 |
| 95 | 94 | 93 | 95 | 93 | 94 | 94 | 93 | 95 | 90 | 92 |
| 100 | 96 | 96 | 96 | 96 | 96 | 96 | 96 | 96 | 96 | 95 |

Abbreviations: Card = Cardiovascular diseases; Mus = Muscoloskeletal diseases; Ment = Mental disorders; M = mean; SD = standard deviation
