## Appendix 5 Descriptive statistics of EPAT-16 sum score per group for "Development and psychometric testing of EPAT-16: A short and valid measure for patient-centeredness from the patient’s perspective"

**Appendix 1: Research checklist: COSMIN Reporting guideline for studies on measurement properties of patient reported outcome measures**

Version August 2021

**Article:** Development and psychometric testing of EPAT-16: A short and valid measure for patient-centeredness from the patient's perspective

Note: The EPAT-16 is not a PROM, but a PREM. Since there are no reporting guidelines for PREMs, we used this guideline where applicable.

| **General Reporting recommendations relevant for all studies on measurement properties** | | | |
|---|---|---|---|
| **Item Number** | **Item Name** | **Item Description** | |
| **Report section: Title** | | | |
| T1 | Patient Reported Outcome Measure (PROM) | The name of the PROM instrument(s) (and version if relevant) being studied | *Title includes "EPAT-16"* |
| T2 | Measurement Property (MP) | What MPs are being studied or more generally, that MPs are being studied (if there are many properties being investigated, for example) | *Title includes "Psychometric testing"* |
| T3 | Study sample | General description of relevant study sample characteristics (e.g., condition of interest, language) and also any intervention or exposure (e.g., treatments) if applicable. | *Our sample were patients in general, which is included in the title* |
| **Report section: Abstract** | | | |
| A1 | PROM | The name of the PROM instrument(s) (and version if relevant) being studied (i.e. the SF-36 or SF-12; language version) or if it concerns an item bank (e.g., PROMIS instruments). The type of instrument (e.g. a self reported questionnaire or interview). | *See Abstract – Background* |
| A2 | Measurement Property | What MPs are being studied or more generally, that MPs are being studied (if there are many properties being investigated, for example) | *Most are mentioned in the methods section of the abstract* |

| | | | |
|---|---|---|---|
| A3 | Design | The type of study being used to test the properties (e.g., test-retest design, longitudinal study, cohort, cross sectional, case series, randomized etc.). Other details of the study design if relevant (intervention/exposure, description of comparison instruments, outcomes other than PROMs). | *See Abstract – Methods* |
| A4 | Sample | Inclusion / exclusion criteria. General description of relevant study sample characteristics (e.g., condition of interest, geographic location, language, other relevant demographic and baseline characteristics) | *See Abstract – Methods* |
| A5 | Methods | A brief description of the methods for investigating each MP including statistical analyses | *See Abstract – Methods* |
| A6 | Results | The main results for all MPs investigated reporting statistics for each result with measures of precision where appropriate. | *See Abstract – Results* |
| A7 | Discussion/Conclusions | A brief description of the results in the context of existing evidence, main strengths and drawbacks and the need for future research on the PROM(s) investigated. | *See Abstract – Conclusions* |
| **Report section: Introduction** | | | |
| I1 | Name and describe the PROM of interest | Specify the name, type, language, and version of the PROM being investigated and how it was developed. Describe the construct the PROM aims to measure and its subscales; describe the structure of the PROM (e.g., the number of factors, the number of items, scoring algorithm); describe relevant instructions (like time period), and number or type of response categories. State whether the PROM is based on a reflective or formative model.<br>Note: This information may also appear in the methods section in greater detail. | *See Background, in particular the first paragraph for EPAT-64 and the last paragraph for EPAT-16* |
| I2 | Target population | Describe the specific target population that the PROM was designed for. The authors need to provide the appropriate and necessary characteristics of this population. | *See Background, last paragraph* |
| I3 | Citation for the original development of the PROM | The citation for the original development paper(s) should be provided and other highly relevant citations related to the quality of the specific PROM under investigation. | *See Background, first paragraph* |

| | | | |
|---|---|---|---|
| I4 | State of Knowledge & Rationale | A description of the current scientific knowledge (what is known) regarding the MPs of the PROM under investigation. The authors should provide a literature review or refer to a recent review of all existing evidence of the specific version (e.g., language, short form) of the PROM and explain why the new study is necessary and important. The rational for the current proposed study should be given. | *Background, second paragraph* |
| I5 | Definitions | Specialized terms should be defined or explained. | *See Background, first paragraph* |
| I6 | Objectives and Hypotheses | State the specific objective(s) of the research and hypotheses related to the specific PROM under investigation. | *See Background, last paragraph* |
| **Report section: General Methods** | | | |
| GM1 | Study Design | State the key elements of the study design | *See Methods, section Data analysis* |
| GM2 | Participants | State how the participants were chosen; the inclusion and exclusion criteria. (e.g., if a PROM for a specific condition, then the eligibility and selection criteria should reflect this). | *See Methods, section Data collection and participants* |
| GM3 | PROM administration | An explicit description of how and when the PROM(s) were administered (e.g., in what setting) including data collection devices/system used (e.g. paper based, electronic administration / ePRO) should be provided. | *See Methods, section Data collection and participants* |
| GM4 | Data collection procedures | Provide information about other data collection, exposure methods (e.g., allocation to interventions) and time points / follow-up points. | *There was no other data collection, exposure to any interventions or further time points.* |
| GM5 | Power/sample size calculation | Provide a power calculation for all MP analyses. Alternatively, if a rule of thumb is used, state it and the source/citation. | *Original sample size calculation was done before data collection and published in a study protocol[1] (see methods - Data collection and participants). Here we describe sample size calculations for all MPs. They were all met by the real sample size of n=1.092 (outpatients) and n=931 (inpatients). Acceptance, relevance, item-difficulty, item-total-correlation and inter-item-correlations: Descriptive statistics/ no power calculation necessary. Confirmatory factor analysis: As a rule of thumb,* |

| | | | |
|---|---|---|---|
| | | | *the COSMIN guidelines recommend a sample size of seven times the number of items (here 16*7=112) and at least 100.*[2]<br>*Construct validity:*<br>*Discriminant validity - To detect a correlation of 0.3 at a significance level of α=0.05 and a test power of 1-β=0.8, a sample of n=85 is necessary.*<br>*Convergent validity: To detect a correlation of 0.5 at a significance level of α=0.05 and a test power of 1-β=0.8, a sample of n=29 is necessary.* |
| GM6 | Statistical analyses | Statistical analyses and tests corresponding to all hypotheses or objectives for all MPs should be reported. Where appropriate, a cut-off for statistical significance should be reported (e.g., p-value less than 0.05). A description of all statistics to be used to estimate the magnitude and direction of effect should also be reported, together with measures of variability or precision. Report statistical package used. | *See Methods, section Data analyses* |
| GM7 | Missing data | State approaches or plan for dealing with missing data. | *See Methods, section Data analyses, last paragraph* |
| GM8 | Post hoc analysis | The report should specify analyses that used data after the data collection period concluded (i.e., if the analyses were post hoc; secondary data analyses) and describe the rationale for any post hoc analyses. | *There were no post hoc or secondary analyses, all analyses were carried out after data collection* |
| **Report section: General Results** | | | |
| GR1 | Missing data | The amount and reasons for missing data should be explained for all analyses for all PROMs (or other outcome measurement instruments) and relevant groups. | *See Table 1 for percentage of missing values per item, we described missing values as proxy for acceptance and know of no other reasons why patients should skip an item* |
| GR2 | Participant/patient Characteristics | The study patients' characteristics should be described, including baseline PROM scores. | *See Appendix 2 for patients' characteristics and Table 1 for PREM scores* |
| GR3 | Sample size | If one study contained analyses using different sample sizes, the authors should report the sample size for each analysis. | *We used the same sample for all analyses* |
| **Report section: Discussion** | | | |

| | | | |
|---|---|---|---|
| D1 | MP evidence | Per measurement property the authors should compare the result to the criteria for good measurement properties (e.g., COSMIN criteria)[27], and determine if the specific MP is sufficient or not. Note: This information may also appear in the results section in greater detail in a table for example. | *See Discussion, section Summary* |
| D2 | Practical relevance | The authors need to discuss the practical relevance of the findings. | *See Discussion, second section* |
| D3 | Strengths and limitations | Strengths and limitations of the study should be discussed. For example, discuss if there were any significant potential biases in the study that could have impacted the results. | *See Discussion, section Limitations and future research* |
| D4 | Generalizability | Generalizability issues related to the PROM results should be discussed. For example, discuss if the results could be generalized to other populations given the sample studied. | *See Discussion, Limitations and future research, last paragraph* |
| D5 | Instrument changes | Discuss the need for modifications to the existing PROM or new PROM development. If you conclude that one of the measurement properties is insufficient, you could suggest some modification, or if it is really poor, you could suggest stopping use of the PROM (in the specific population or in general). | *See Discussion, Limitations and future research, last paragraph* |
| D6 | Future Research | Report specifically the type of research needed to answer new questions arising out of these findings for the particular MP and PROM investigated. | *See Discussion, Limitations and future research, last paragraph* |
| **Report section: Conclusions** | | | |
| C1 | Conclusions | State the overall conclusions for each MP and of the use PROM investigated. | *See Discussion, section Conclusion* |
| **Report section: Other information** | | | |
| O1 | Conflict of Interest | State any relevant conflict of interest related to the PROM under investigation (e.g., an author being the PROM developer, funding body etc). | *See Competing interests* |

| **Specific Reporting recommendations for studies on Structural Validity** | | | |
|---|---|---|---|
| **Item Number** | **Item Name** | **Item Description** | |
| SV1 | Factor Analyses: Classical | Report details of the methods and results for any exploratory or | *See Methods, section Data analysis, p. 5, ll. 23-23* |

| | | | |
|---|---|---|---|
| | Test Theory (CTT) PROMs | confirmatory factor analyses. State the rational for any explorative factor analyses (e.g., no clear a priori hypotheses). For CFA, describe and justify the factor structure of tested models. Methods and results for checking of the assumptions should be described, the method of estimation, goodness-of-fit statistics and cut-off points for good model fit, including factor loadings of best-fitting model. | *and Appendix 2* |
| SV2 | Item Response Theory (IRT) analyses | Type of IRT/Rasch model should be reported. Also report the method of estimation, methods and results for checking of the assumptions (unidimensionality (see factor analysis), local dependency (e.g., residual correlations), monotonicity; (e.g. Mokken scaling), goodness-of-fit statistics, and cut-off points for goodness of item/model fit, and all item parameters. | *Not applicable* |

| **Specific Reporting recommendations for studies on Internal Consistency** | | | |
|---|---|---|---|
| **Item Number** | **Item Name** | **Item Description** | |
| IC1 | Unit of measurement | Report internal consistency methods and results for each unidimensional scale or subscale. Report all evidence or assumptions associated with unidimensionality. | *See Methods, subsection Psychometric analyses of final EPAT-16, first paragraph* |
| IC2 | Continuous scores | Report Cronbach's alpha or omega statistics. Report other statistics calculated for internal consistency of continuous scores. | *See results, subsection Psychometric analyses of final EPAT-16* |
| IC3 | Dichotomous scores | Report Cronbach's alpha or Kuder-Richardson coefficient. Report other statistics calculated for internal consistency of dichotomous scores. | *Not applicable* |

| **Specific Reporting recommendations for studies on Hypotheses Testing for Construct Validity** | | | |
|---|---|---|---|
| **Item Number** | **Item Name** | **Item Description** | |
| ConV1 | Comparator instrument(s) | The comparator instruments should be appropriately described in terms of the construct(s) they intend to measure. Report the measurement properties of the comparator instruments and related citations or data. | *See Methods, subsection Psychometric analyses of final EPAT-16, second paragraph* |

| | | | |
|---|---|---|---|
| ConV2 | Comparator Group(s) | Report characteristics of groups being compared. Include sample sizes in each group. | *Not applicable* |
| ConV3 | Hypotheses | Report all hypotheses including the direction and magnitude of the expected correlations between the PROM of interest and another measurement instrument, or the direction and magnitude of differences in scores of the PROM between groups. | *See Methods, subsection Psychometric analyses of final EPAT-16, second paragraph* |
| ConV4 | Statistical analyses | Report all statistical methods and results used to test each hypothesis. | *See Results, subsection Psychometric analyses of final EPAT-16, second paragraph* |
| ConV5 | Results | Report which specific results are in accordance with its hypothesis. | *See Results, subsection Psychometric analyses of final EPAT-16, second paragraph* |

| **Specific Reporting recommendations for studies on Cross-Cultural Validity\Measurement Invariance** | | | |
|---|---|---|---|
| **Item Number** | **Item Name** | **Item Description** | |
| CCV1 | Comparator Group(s) | Report characteristics of (sub)groups being compared. Include sample sizes in each group. | See Appendix 2 |
| CCV2 | Factor Analyses: Classical Test Theory (CTT) PROMs | Report details of the methods and results for multiple-group confirmatory factor analyses, logistic regression analyses, or other analyses performed. Describe and justify the series of tested models, including constraints of factor loadings, intercepts and variances in CFA. Methods and results for checking of the assumptions should be described. criteria to define invariance. Describe the method of estimation, goodness-of-fit statistics and criteria used to flag items for measurement invariance. | See Methods, subsection Psychometric analyses of final EPAT-16, first paragraph |
| CCV3 | Item Response Theory (IRT) analyses | Type of IRT/Rasch model should be reported. Also report the methods and results for checking of the assumptions (unidimensionality (see factor analysis), local dependency (e.g., residual correlations), monotonicity; (e.g. Mokken scaling).. Describe statistical packages, method of estimation, criteria used to flag items for DIF, and methods and results of all model comparisons. | Not applicable |
